# QRS fragmentation as a predictor of clinical events in patients undergoing cardiac resynchronization therapy

**DOI:** 10.1101/2025.04.01.25325064

**Authors:** Carlos Rodríguez López, Lara de Miguel García, Jorge Balaguer Germán, Marcelino Cortés García, Álvaro Aceña Navarro, Camila Sofía García Talavera, José María Romero Otero, Carla Rivero Lázaro, Loreto Bravo Calero, Francisco Díaz Cortegana, José Tuñón, José-Manuel Rubio Campal

## Abstract

**Background:** QRS fragmentation (QRSf) has been associated with a worse prognosis in several cardiac diseases, although limited evidence exists on QRSf in patients undergoing cardiac resynchronization therapy (CRT).

**Objectives:** This study aimed to determine whether QRSf is associated with clinical events in patients receiving CRT.

**Methods:** QRSf included various patterns present on at least 2 contiguous leads in 3 different territories: anteroseptal (V1-V4), lateral (V5-V6, I, aVL), and inferior (II, III, aVF). Mortality, heart failure (HF) admissions, and arrhythmic events (AEs) were studied.

**Results:** We included 244 patients (mean age, 72 ± 10 years; 80% male). CRT with a defibrillator (CRT-D) was implanted in 176 patients. Before implantation, mean QRS width was 158 ± 29 ms, mean LVEF was 25 ± 9%, and 61% had QRSf. With pacing, new QRSf developed in 11.9% of patients and QRSf resolved in 11.5%. After a median follow-up of 36 months, 36.9% of patients died, 37.3% were hospitalized for HF, and 16.5% of those with CRT-D had an AE. On multivariate analysis, pre-implantation QRSf was an independent predictor of all-cause mortality (hazard ratio (HR), 2.79; 95% CI [1.54-5.05]), hospitalization for HF (HR, 3.57; 95% CI [1.94-6.58]), and AEs (HR, 6.99; 95% CI [1.54-31.68]). Patients with persistent or newly developed QRSf after implantation had a worse prognosis than those without QRSf.

**Conclusions:** QRSf before or after CRT has significant prognostic value for all-cause mortality, HF hospitalization, and AEs, and presence of QRSf after implantation also has prognostic significance. Patients with QRSf before or after CRT should be observed more closely, as they have a worse prognosis.

## INTRODUCTION

Cardiac resynchronization therapy (CRT), with implantable cardioverter defibrillator (ICD; CRT-D) or without ICD (CRT-P), has been shown to reduce mortality and hospitalization in patients with heart failure (HF) and improve their quality of life (1,2). However, as HF encompasses a highly varied group of patients, a significant percentage of non-responders have a worse prognosis at long-term follow-up (3,4). Therefore, there is a need to identify new features of HF to help stratify risk, optimize prognosis, and predict treatment response.

In recent years, studies have shown that QRS fragmentation (QRSf) is a potential prognostic marker in several cardiac diseases (5–8). However, the evidence in support of using this ECG finding in patients undergoing CRT is not robust, as it is based on small studies with limited sample sizes (9–11). Furthermore, the evolution of QRSf after device implantation and its significance have not been fully described. We present an observational study of a large cohort of patients undergoing CRT in which we assess the prognostic implications of QRSf before and after cardiac pacing implantation.

## METHODS

### Study design

We conducted a retrospective, single-center, observational study of consecutive patients who underwent CRT/CRT-D in the cardiology department of Fundación Jiménez Díaz University Hospital (Madrid, Spain). This study was approved by the research ethics committee of our institution (EO284-23) and conducted in accordance with the guidelines of the Declaration of Helsinki.

### Study population and protocol

Patients were recruited between October 2009 and December 2022. All participants underwent device implantation in accordance with current ESC and AHA-HRS guidelines (1,3,12) and received optimal medical therapy when appropriate. **Patients with baseline narrow QRS complexes were included if CRT was implanted because of an indication for pacing for another reason.** The most significant exclusion criteria were device explantation, failure to obtain a pre- or post-implantation ECG, severe valvular heart disease with an indication for intervention, cardiac transplantation, and left ventricular assist device implantation (Figure S1). Eligible patients underwent a 12-lead surface ECG, blood sampling and a comprehensive transthoracic echocardiography before CRT. At the end of follow-up, the occurrence of the primary and secondary endpoints was assessed either by review of electronic medical records (Figure S2).

### Electrocardiogram analysis

A resting 12-lead surface ECG (TraceMaster, Philips, filter range, 0.05-150 Hz; 25 mm/s; 10 mm/mV) was obtained for each participant at least 24 hours before and one month after CRT implantation. QRSf was defined in accordance with the criteria of Das et al (5,6). For QRS complexes ≥120 ms, fQRS was defined by the presence of various RSR patterns, patterns with or without a Q wave, with >2 R waves (R’) or >2 notches in the R wave, or in the downstroke or upstroke of the S wave, in 2 contiguous leads corresponding to a major coronary artery territory. For paced QRS complexes, fQRS was defined by the presence of >2 R’ or >2 notches in the S waves in 2 contiguous leads. For QRS complexes <120 ms, fragmentation was identified in the presence of an additional R’ or notching of the R or S waves, or the presence of more than two R’ in two contiguous leads. As shown in Figure 1, the location of the QRSf was distributed over 3 territories: anteroseptal (V1-V4), lateral (V5-V6, I, aVL), and inferior (II, III, and aVF). When QRSf was observed in more than one region, the location was assigned based on the number of leads showing QRSf in each territory. **The resolution of QRSf was defined as occurring when a patient who had previously exhibited QRSf did not meet the criteria for fragmentation in the CRT-paced QRS complex**. **The variation of the location of QRSf between baseline QRS complex and CRT-paced QRS complex was not considered as resolution of QRSf.** QRS pre- and post-implantation ECGs were interpreted independently by two experienced cardiologists blinded to the clinical variables and results. In cases of disagreement, the final decision was made by a third experienced cardiologist.

**Figure 1.**
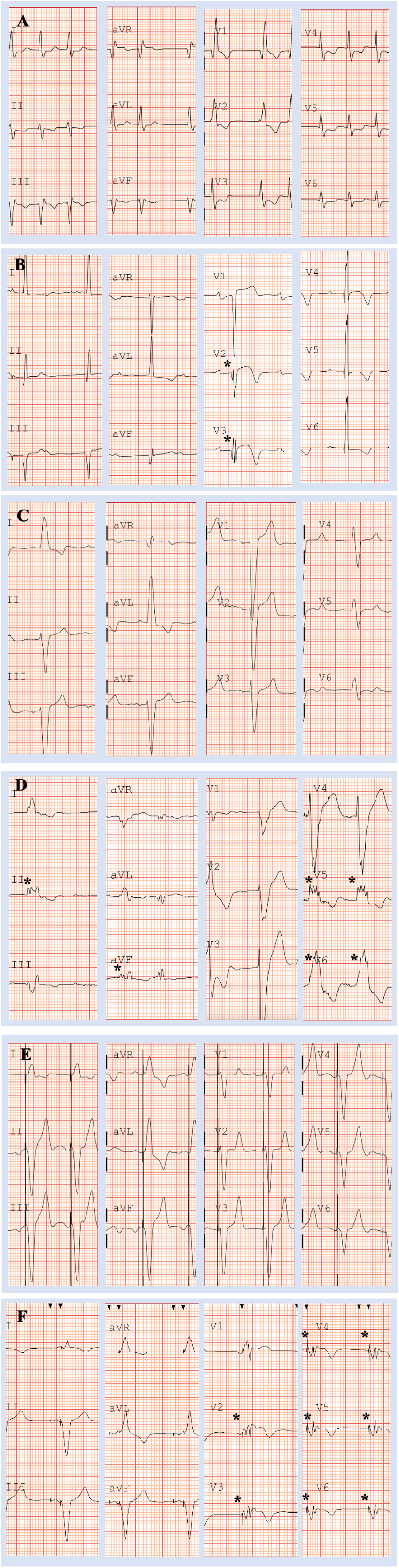
Fragmented and non-fragmented QRS complexes. **A.** Non-fragmented ECG with RBBB morphology. **B.** Fragmented narrow QRS. **C.** Non-fragmented QRS with LBBB morphology. **D.** Fragmented QRS with LBBB morphology in inferior and lateral leads. **D.** Paced non-fragmented ECG. **E.** Paced fragmented QRS in anterior and lateral leads.

### Transthoracic echocardiography

At least one month before the start of CRT and annually during follow-up, transthoracic echocardiography (TTE) was performed by experienced sonographers using a commercially available system (EPIQ CVx 7.0.3, Philips Healthcare, Best, The Netherlands) equipped with an X5–1 xMATRIX array transducer, following a standardized protocol. Left ventricular ejection fraction (LVEF), LV end-diastolic dimension (LVEDD), and left atrial anteroposterior diameter (LAAPD) were assessed according to guidelines of the American Society of Echocardiography and the European Association of Cardiovascular Imaging (13,14). Severity of valvulopathies was categorized as absent or trivial, mild, moderate, or severe, according to guidelines recommendations (15,16).

### CRT and CRT-D implantation, programming, optimization, and interrogation

Commercially available transvenous CRT with or without defibrillator devices were implanted according to standardized protocol. The coronary sinus lead was placed in a lateral, posterolateral, or posterior position whenever possible, with an anterior position reserved only as a last resort. The left ventricular lead position was retrospectively evaluated using left anterior oblique and right anterior oblique views by an experienced electrophysiologist.

CRT devices were programmed to achieve the narrowest QRS complex using specific algorithms (2). We used specific vendors device-based algorithms for optimization of atrioventricular (AV) and interventricular (VV) delay, LV-only pacing and fusion pacing to maximize the reduction of the QRS duration including Sync-AV, QuickOpt, multipoint pacing (MPP), Adaptative CRT, Smart Delay and aCRT. For patients where the algorithms could not be used, typical programming included a fixed AV time at least 20 ms shorter than the intrinsic conduction time to ensure biventricular capture and simultaneous pacing of the right and left ventricle, or (in patients with permanent atrial fibrillation) synchronous biventricular pacing without trigger mode.

CRT-Ds were programmed to deliver a shock for ventricular rates between 181 and 188 bpm and antitachycardia pacing between 150 and 181 bpm. Appropriate CRT-D therapies were reviewed during follow-up by experienced electrophysiologists blinded to ECG findings. Recorded electrograms from episodes of CRT-D therapy were available for this study. An arrhythmic event (AE) was defined as delivery of appropriate ICD therapy (antitachycardia pacing or ICD shock) for ventricular arrhythmia or sustained ventricular arrhythmia not requiring ICD therapy. Inappropriate ICD therapy was also determined by reviewing saved episode-specific electrograms and was not included as an AE.

### Endpoints

The primary outcome was the association between the presence of QRSf before or after implantation and all-cause mortality or hospitalization for HF, defined as admission to a health care facility for more than 24 hours with symptoms of congestive HF and subsequent treatment. Admissions for other medical conditions that subsequently developed into HF were not classified as hospitalizations. Secondary endpoints included the association between AEs and their correlation with clinical variables. Mortality data were obtained by chart review and by searching websites made available by the Spanish public health system.

### Statistical analysis

All continuous variables following a normal distribution are reported as mean ± standard deviation (SD). Non-normal variables as determined by Kolmogorov-Smirnoff and Saphiro-Wilk normality tests are presented as median (interquartile range [IQR]). Unpaired *t* tests were used to compare quantitative variables across groups. When comparing continuous variables across multiple groups, variance analysis was performed. Discrete and categorical variables are presented as frequencies and percentages. To compare categorical variables between groups, *χ*^2^ or Fisher’s exact test was performed. Univariate Cox regression analysis was used to determine hazard ratios (HRs) and 95% confidence interval (CI) of different variables with outcomes. Then, each variable with a significance <0.2 was included as a covariate in a multivariate Cox regression analysis. Stepwise selection with a *P*-value of 0.05 for backward selection was used to select the most strongly associated variables. Cumulative event-free survival curves were constructed using Kaplan-Meier analysis with log-rank testing for comparison. All statistical tests were two-tailed with *P*-values <0.05 indicating statistical significance. The SPSS software package, version 27.0, was used for all analyses. **For evaluating the interobserver and intraobserver concordance and agreement between QRSf status, Cohen’s kappa was calculated.**

## RESULTS

A total of 244 patients (mean age, 72 ± 10 years; 80% male) were included. Baseline characteristics are shown in Table 1 and Table S1. A CRT-D was implanted in 176 patients (72.2%). At baseline, 155 patients had QRSf (19% anteroseptal, 12% lateral, and 30% inferior). After CRT implantation, 29 patients (11.9%) without prior QRSf developed QRSf, whereas 40 patients (16.5%) experienced QRSf resolution. The distribution of QRSf after implantation was as follows: anteroseptal 25%, lateral 13%, and inferior 19%. The mean pre-implantation QRS width was 159 ± 29 ms, and 79% of patients had left bundle branch block (LBBB). Patients with QRSf had wider QRS complexes and more frequent LBBB. The mean paced QRS width was 141 ± 23 ms. Mean LVEF before implantation was 25 ± 9 %, mean LVEDD was 56.5 ± 9 mm, and mean LAAPD was 44 ± 9 mm. Seventy-six patients (31.7%) had moderate to severe mitral regurgitation at baseline. Patients with QRSf either before or after device implantation had significantly lower LVEF and larger LVEDD at baseline (Table 1). The median follow-up time for patients receiving CRT was 36.0 months (IQR, 18.2-75.7), while the median time of follow-up in patients undergoing CRT-D was 43.6 months (IQR, 19.5-81.1).

**Table 1.**
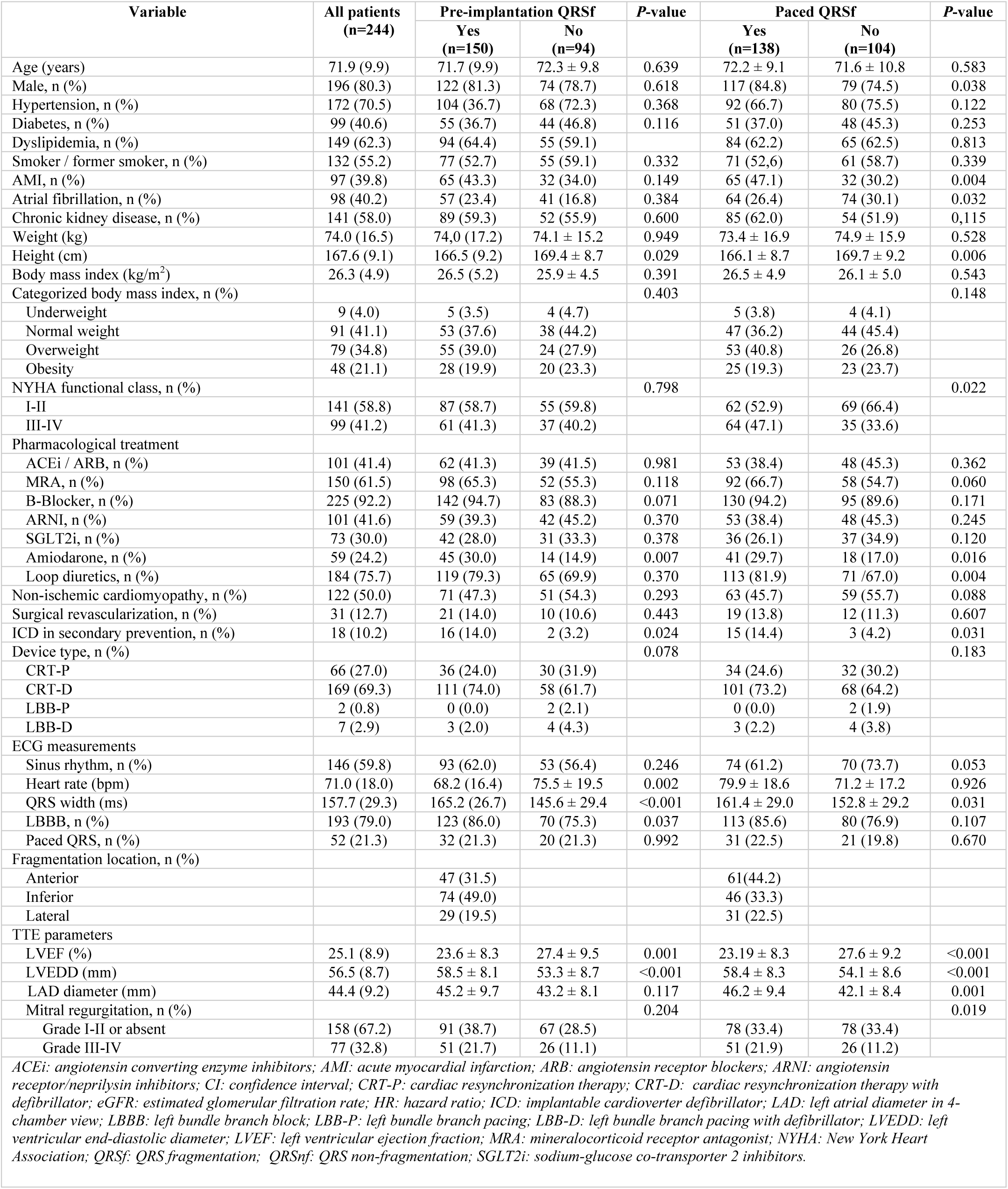
Baseline characteristics (n = 244)

| Variable | All patients<br>(n=244) | Pre-implantation QRSf |  | P-value | Paced QRSf |  | P-value |
| --- | --- | --- | --- | --- | --- | --- | --- |
|  |  | Yes<br>(n=150) | No<br>(n=94) |  | Yes<br>(n=138) | No<br>(n=104) |  |
| Age (years) | 71.9 (9.9) | 71.7 (9.9) | 72.3 ± 9.8 | 0.639 | 72.2 ± 9.1 | 71.6 ± 10.8 | 0.583 |
| Male, n (%) | 196 (80.3) | 122 (81.3) | 74 (78.7) | 0.618 | 117 (84.8) | 79 (74.5) | 0.038 |
| Hypertension, n (%) | 172 (70.5) | 104 (36.7) | 68 (72.3) | 0.368 | 92 (66.7) | 80 (75.5) | 0.122 |
| Diabetes, n (%) | 99 (40.6) | 55 (36.7) | 44 (46.8) | 0.116 | 51 (37.0) | 48 (45.3) | 0.253 |
| Dyslipidemia, n (%) | 149 (62.3) | 94 (64.4) | 55 (59.1) |  | 84 (62.2) | 65 (62.5) | 0.813 |
| Smoker / former smoker, n (%) | 132 (55.2) | 77 (52.7) | 55 (59.1) | 0.332 | 71 (52.6) | 61 (58.7) | 0.339 |
| AMI, n (%) | 97 (39.8) | 65 (43.3) | 32 (34.0) | 0.149 | 65 (47.1) | 32 (30.2) | 0.004 |
| Atrial fibrillation, n (%) | 98 (40.2) | 57 (23.4) | 41 (16.8) | 0.384 | 64 (26.4) | 74 (30.1) | 0.032 |
| Chronic kidney disease, n (%) | 141 (58.0) | 89 (59.3) | 52 (55.9) | 0.600 | 85 (62.0) | 54 (51.9) | 0.115 |
| Weight (kg) | 74.0 (16.5) | 74.0 (17.2) | 74.1 ± 15.2 | 0.949 | 73.4 ± 16.9 | 74.9 ± 15.9 | 0.528 |
| Height (cm) | 167.6 (9.1) | 166.5 (9.2) | 169.4 ± 8.7 | 0.029 | 166.1 ± 8.7 | 169.7 ± 9.2 | 0.006 |
| Body mass index (kg/m<sup>2</sup>) | 26.3 (4.9) | 26.5 (5.2) | 25.9 ± 4.5 | 0.391 | 26.5 ± 4.9 | 26.1 ± 5.0 | 0.543 |
| Categorized body mass index, n (%) |  |  |  | 0.403 |  |  | 0.148 |
| Underweight | 9 (4.0) | 5 (3.5) | 4 (4.7) |  | 5 (3.8) | 4 (4.1) |  |
| Normal weight | 91 (41.1) | 53 (37.6) | 38 (44.2) |  | 47 (36.2) | 44 (45.4) |  |
| Overweight | 79 (34.8) | 55 (39.0) | 24 (27.9) |  | 53 (40.8) | 26 (26.8) |  |
| Obesity | 48 (21.1) | 28 (19.9) | 20 (23.3) |  | 25 (19.3) | 23 (23.7) |  |
| NYHA functional class, n (%) |  |  |  | 0.798 |  |  | 0.022 |
| I-II | 141 (58.8) | 87 (58.7) | 55 (59.8) |  | 62 (52.9) | 69 (66.4) |  |
| III-IV | 99 (41.2) | 61 (41.3) | 37 (40.2) |  | 64 (47.1) | 35 (33.6) |  |
| Pharmacological treatment |  |  |  |  |  |  |  |
| ACEi / ARB, n (%) | 101 (41.4) | 62 (41.3) | 39 (41.5) | 0.981 | 53 (38.4) | 48 (45.3) | 0.362 |
| MRA, n (%) | 150 (61.5) | 98 (65.3) | 52 (55.3) | 0.118 | 92 (66.7) | 58 (54.7) | 0.060 |
| B-Blocker, n (%) | 225 (92.2) | 142 (94.7) | 83 (88.3) | 0.071 | 130 (94.2) | 95 (89.6) | 0.171 |
| ARNI, n (%) | 101 (41.6) | 59 (39.3) | 42 (45.2) | 0.370 | 53 (38.4) | 48 (45.3) | 0.245 |
| SGLT2i, n (%) | 73 (30.0) | 42 (28.0) | 31 (33.3) | 0.378 | 36 (26.1) | 37 (34.9) | 0.120 |
| Amiodarone, n (%) | 59 (24.2) | 45 (30.0) | 14 (14.9) | 0.007 | 41 (29.7) | 18 (17.0) | 0.016 |
| Loop diuretics, n (%) | 184 (75.7) | 119 (79.3) | 65 (69.9) | 0.370 | 113 (81.9) | 71 (67.0) | 0.004 |
| Non-ischemic cardiomyopathy, n (%) | 122 (50.0) | 71 (47.3) | 51 (54.3) | 0.293 | 63 (45.7) | 59 (55.7) | 0.088 |
| Surgical revascularization, n (%) | 31 (12.7) | 21 (14.0) | 10 (10.6) | 0.443 | 19 (13.8) | 12 (11.3) | 0.607 |
| ICD in secondary prevention, n (%) | 18 (10.2) | 16 (14.0) | 2 (3.2) | 0.024 | 15 (14.4) | 3 (4.2) | 0.031 |
| Device type, n (%) |  |  |  | 0.078 |  |  | 0.183 |
| CRT-P | 66 (27.0) | 36 (24.0) | 30 (31.9) |  | 34 (24.6) | 32 (30.2) |  |
| CRT-D | 169 (69.3) | 111 (74.0) | 58 (61.7) |  | 101 (73.2) | 68 (64.2) |  |
| LBB-P | 2 (0.8) | 0 (0.0) | 2 (2.1) |  | 0 (0.0) | 2 (1.9) |  |
| LBB-D | 7 (2.9) | 3 (2.0) | 4 (4.3) |  | 3 (2.2) | 4 (3.8) |  |
| ECG measurements |  |  |  |  |  |  |  |
| Sinus rhythm, n (%) | 146 (59.8) | 93 (62.0) | 53 (56.4) | 0.246 | 74 (61.2) | 70 (73.7) | 0.053 |
| Heart rate (bpm) | 71.0 (18.0) | 68.2 (16.4) | 75.5 ± 19.5 | 0.002 | 79.9 ± 18.6 | 71.2 ± 17.2 | 0.926 |
| QRS width (ms) | 157.7 (29.3) | 165.2 (26.7) | 145.6 ± 29.4 | <0.001 | 161.4 ± 29.0 | 152.8 ± 29.2 | 0.031 |
| LBBB, n (%) | 193 (79.0) | 123 (86.0) | 70 (75.3) | 0.037 | 113 (85.6) | 80 (76.9) | 0.107 |
| Paced QRS, n (%) | 52 (21.3) | 32 (21.3) | 20 (21.3) | 0.992 | 31 (22.5) | 21 (19.8) | 0.670 |
| Fragmentation location, n (%) |  |  |  |  |  |  |  |
| Anterior |  | 47 (31.5) |  |  | 61 (44.2) |  |  |
| Inferior |  | 74 (49.0) |  |  | 46 (33.3) |  |  |
| Lateral |  | 29 (19.5) |  |  | 31 (22.5) |  |  |
| TTE parameters |  |  |  |  |  |  |  |
| LVEF (%) | 25.1 (8.9) | 23.6 ± 8.3 | 27.4 ± 9.5 | 0.001 | 23.19 ± 8.3 | 27.6 ± 9.2 | <0.001 |
| LVEDD (mm) | 56.5 (8.7) | 58.5 ± 8.1 | 53.3 ± 8.7 | <0.001 | 58.4 ± 8.3 | 54.1 ± 8.6 | <0.001 |
| LAD diameter (mm) | 44.4 (9.2) | 45.2 ± 9.7 | 43.2 ± 8.1 | 0.117 | 46.2 ± 9.4 | 42.1 ± 8.4 | 0.001 |
| Mitral regurgitation, n (%) |  |  |  | 0.204 |  |  | 0.019 |
| Grade I-II or absent | 158 (67.2) | 91 (38.7) | 67 (28.5) |  | 78 (33.4) | 78 (33.4) |  |
| Grade III-IV | 77 (32.8) | 51 (21.7) | 26 (11.1) |  | 51 (21.9) | 26 (11.2) |  |

ACEi: angiotensin converting enzyme inhibitors; AMI: acute myocardial infarction; ARB: angiotensin receptor blockers; ARNI: angiotensin receptor/neprilysin inhibitors; CI: confidence interval; CRT-P: cardiac resynchronization therapy; CRT-D: cardiac resynchronization therapy with defibrillator; eGFR: estimated glomerular filtration rate; HR: hazard ratio; ICD: implantable cardioverter defibrillator; LAD: left atrial diameter in 4-chamber view; LBBB: left bundle branch block; LBB-P: left bundle branch pacing; LBB-D: left bundle branch pacing with defibrillator; LVEDD: left ventricular end-diastolic diameter; LVEF: left ventricular ejection fraction; MRA: mineralocorticoid receptor antagonist; NYHA: New York Heart Association; QRSf: QRS fragmentation; QRSnf: QRS non-fragmentation; SGLT2i: sodium-glucose co-transporter 2 inhibitors.

At follow-up, 90 patients had died, 91 had at least one hospitalization for an HF-related event, and 29 participants who received CRT-D had an AE (Tables 2-3). The mean LVEF was 33 ± 14% and the mean LVEDD was 54 ± 9 mm. Patients with QRSf either before or after CRT implantation had significantly lower LVEF and greater LVEDD compared to patients without QRSf. Significant differences in LVEF and LVEDD were also found between subgroups according to pre- and post-QRSf status. LVEF improved by at least 10% in 89 patients and LVEDD improved in 113 patients. One hundred eighty-six patients were in New York Heart Association (NYHA) functional class I-II at follow-up, while 103 had an improvement in NYHA functional class. A significant difference in the proportion of patients with LVEF improvement over 10% and NYHA functional class I or II at follow-up was observed between patients with and without paced QRSf (Table 2).

**Table 2.**
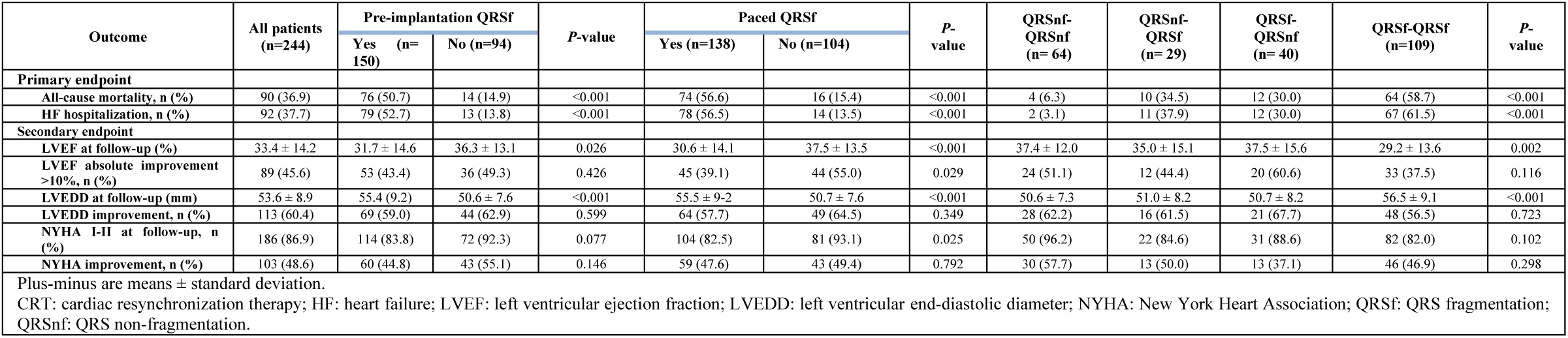
Primary and secondary outcomes in patients undergoing CRT.

| Outcome | All patients<br>(n=244) | Pre-implantation QRSf |  | P-value | Paced QRSf |  | P-value | QRSnf-QRSnf<br>(n= 64) | QRSnf-QRSf<br>(n= 29) | QRSf-QRSnf<br>(n= 40) | QRSf-QRSf<br>(n=109) | P-value |
| --- | --- | --- | --- | --- | --- | --- | --- | --- | --- | --- | --- | --- |
|  |  | Yes (n=150) | No (n=94) |  | Yes (n=138) | No (n=104) |  |  |  |  |  |  |
| **Primary endpoint** |  |  |  |  |  |  |  |  |  |  |  |  |
| All-cause mortality, n (%) | 90 (36.9) | 76 (50.7) | 14 (14.9) | <0.001 | 74 (56.6) | 16 (15.4) | <0.001 | 4 (6.3) | 10 (34.5) | 12 (30.0) | 64 (58.7) | <0.001 |
| HF hospitalization, n (%) | 92 (37.7) | 79 (52.7) | 13 (13.8) | <0.001 | 78 (56.5) | 14 (13.5) | <0.001 | 2 (3.1) | 11 (37.9) | 12 (30.0) | 67 (61.5) | <0.001 |
| **Secondary endpoint** |  |  |  |  |  |  |  |  |  |  |  |  |
| LVEF at follow-up (%) | 33.4 ± 14.2 | 31.7 ± 14.6 | 36.3 ± 13.1 | 0.026 | 30.6 ± 14.1 | 37.5 ± 13.5 | <0.001 | 37.4 ± 12.0 | 35.0 ± 15.1 | 37.5 ± 15.6 | 29.2 ± 13.6 | 0.002 |
| LVEF absolute improvement >10%, n (%) | 89 (45.6) | 53 (43.4) | 36 (49.3) | 0.426 | 45 (39.1) | 44 (55.0) | 0.029 | 24 (51.1) | 12 (44.4) | 20 (60.6) | 33 (37.5) | 0.116 |
| LVEDD at follow-up (mm) | 53.6 ± 8.9 | 55.4 (9.2) | 50.6 ± 7.6 | <0.001 | 55.5 ± 9.2 | 50.7 ± 7.6 | <0.001 | 50.6 ± 7.3 | 51.0 ± 8.2 | 50.7 ± 8.2 | 56.5 ± 9.1 | <0.001 |
| LVEDD improvement, n (%) | 113 (60.4) | 69 (59.0) | 44 (62.9) | 0.599 | 64 (57.7) | 49 (64.5) | 0.349 | 28 (62.2) | 16 (61.5) | 21 (67.7) | 48 (56.5) | 0.723 |
| NYHA I-II at follow-up, n (%) | 186 (86.9) | 114 (83.8) | 72 (92.3) | 0.077 | 104 (82.5) | 81 (93.1) | 0.025 | 50 (96.2) | 22 (84.6) | 31 (88.6) | 82 (82.0) | 0.102 |
| NYHA improvement, n (%) | 103 (48.6) | 60 (44.8) | 43 (55.1) | 0.146 | 59 (47.6) | 43 (49.4) | 0.792 | 30 (57.7) | 13 (50.0) | 13 (37.1) | 46 (46.9) | 0.298 |

Plus-minus are means ± standard deviation. CRT: cardiac resynchronization therapy; HF: heart failure; LVEF: left ventricular ejection fraction; LVEDD: left ventricular end-diastolic diameter; NYHA: New York Heart Association; QRSf: QRS fragmentation; QRSnf: QRS non-fragmentation.

**Table 3.** Arrhythmic events in patients undergoing CRT-D.

| Table 3. Arrhythmic events in patients undergoing CRT-D. |  |  |  |  |  |  |  |  |  |  |  |  |
| --- | --- | --- | --- | --- | --- | --- | --- | --- | --- | --- | --- | --- |
| Outcome, n (%) | All patients<br>(n=176) | Pre-implantation QRSf |  | P-value | Paced QRSf |  | P-value | QRSnf-QRSnf<br>(n=44) | QRSnf-QRSf<br>(n=17) | QRSf-QRSnf<br>CRT (n=26) | QRSf-QRSf<br>(n=87) | P-value |
|  |  | Yes<br>(n=114) | No (n=62) |  | Yes<br>(n=70) | No<br>(n=104) |  |  |  |  |  |  |
| Secondary endpoint |  |  |  |  |  |  |  |  |  |  |  |  |
| Sustained VT or appropriate ICD therapies | 29 (16.5) | 27 (23.7) | 2 (3.2) | <0.001 | 23 (22.1) | 6 (8.6) | 0.019 | 3 (6.8) | 0 (0.0) | 3 (11.5) | 23 (26.4) | 0.005 |

CRT-D: cardiac resynchronization therapy with defibrillator; ICD: implantable cardioverter defibrillator; QRSf: QRS fragmentation; QRSnf: QRS non-fragmentation; VT: ventricular tachycardia.

### All-cause mortality

Ninety patients (36.9%) died during follow-up. Among them, 76 had QRSf before implantation and 74 had paced QRSf after implantation. Four of 64 patients with no QRSf before or after CRT died, whereas 64 of 109 patients with QRSf before and after CRT died during follow-up. A total of 10 of 29 patients who developed paced QRSf after CRT died, whereas 12 of 28 patients in whom QRSf was corrected after CRT pacing died during follow-up (Table 2).

On univariate analysis using Cox regression (Table 4), all-cause mortality was significantly associated with age, female sex, non-ischemic etiology, use of loop diuretics, pre-implantation QRSf, paced QRS width, and LVEDD and LAAPD at follow-up. A multivariate analysis using Cox regression revealed that all-cause mortality remained significantly associated with baseline QRSf and age. Baseline QRSf was strongly associated with this primary outcome (HR 2.79; 95% CI [1.54-5.05]). In addition, a multivariate analysis including the presence of paced QRSf after CRT implantation compared to native QRSf showed that paced QRSf after CRT implantation was also independently associated with all-cause mortality (Table S2). These two variables were not included in the same multivariate analysis because they appeared to be strongly related, which could introduce high collinearity. Kaplan-Meier curves showed significant differences in survival between patients with and without pre-implantation QRSf and paced QRSf (Figures 2A and 2B. When categorizing the pre- and post-implantation QRSf status (QRSf and QRSf, QRSf and non-QRSf, non-QRSf and QRSf, and non-QRSf and non-QRSf), the multivariate Cox regression survival analysis showed that the presence of QRSf before implantation, with or without paced QRSf after CRT, was independently associated with all-cause mortality (HR, 4.43; 95% CI [1.53-12.8] and HR, 3.69; 95% CI [1.16-11.8], respectively). Participants without QRSf prior to implantation who developed QRSf after CRT did not have a significantly higher risk of all-cause mortality (Table S3). Kaplan-Meier curves according to QRSf status before and after CRT are presented in Figure 2C.

**Table 4.**
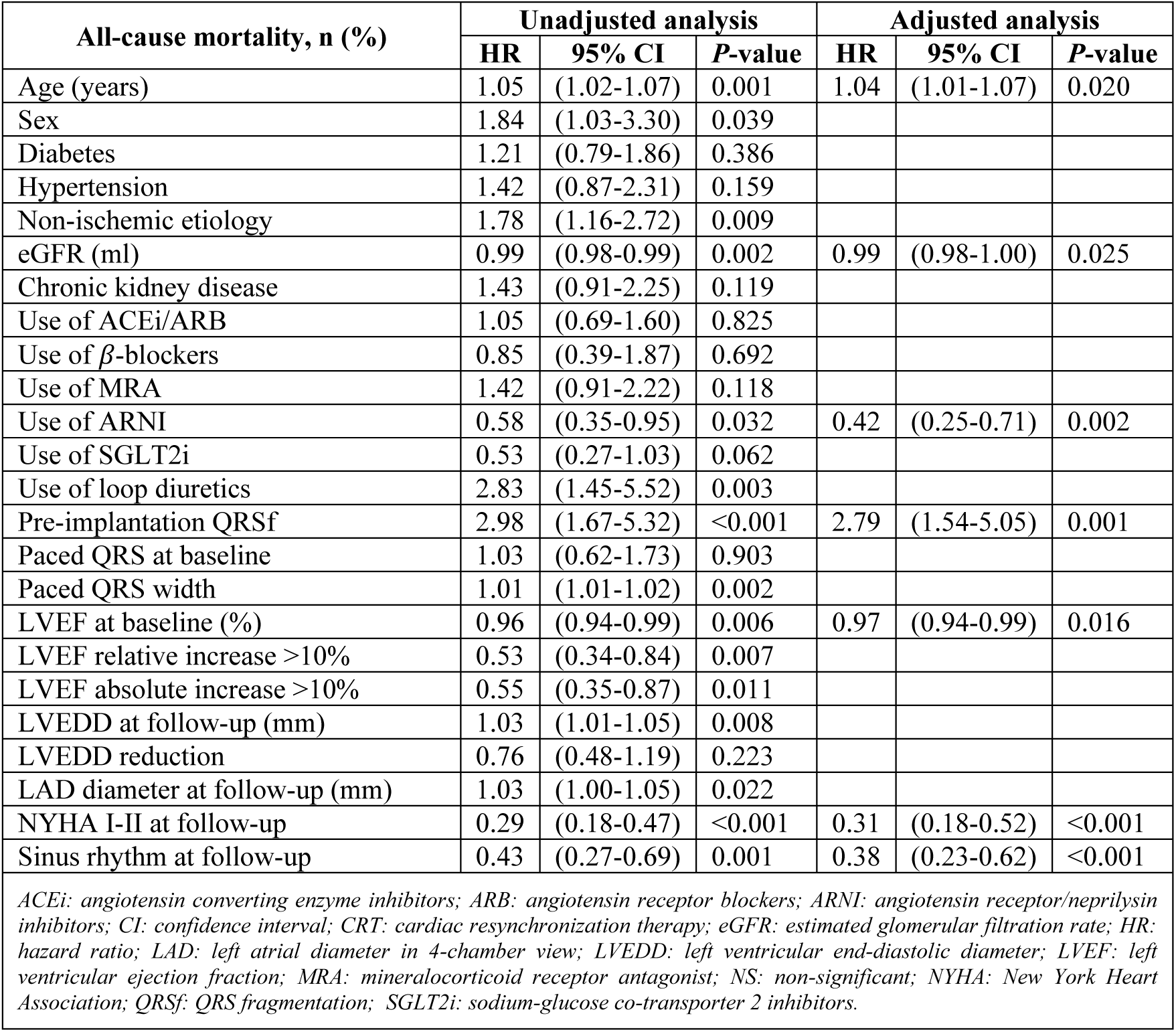
Univariate and multivariate analysis using Cox regression for all-cause mortality according to pre-implantation QRSf.

| All-cause mortality, n (%) | Unadjusted analysis |  |  | Adjusted analysis |  |  |
| --- | --- | --- | --- | --- | --- | --- |
|  | HR | 95% CI | P-value | HR | 95% CI | P-value |
| Age (years) | 1.05 | (1.02-1.07) | 0.001 | 1.04 | (1.01-1.07) | 0.020 |
| Sex | 1.84 | (1.03-3.30) | 0.039 |  |  |  |
| Diabetes | 1.21 | (0.79-1.86) | 0.386 |  |  |  |
| Hypertension | 1.42 | (0.87-2.31) | 0.159 |  |  |  |
| Non-ischemic etiology | 1.78 | (1.16-2.72) | 0.009 |  |  |  |
| eGFR (ml) | 0.99 | (0.98-0.99) | 0.002 | 0.99 | (0.98-1.00) | 0.025 |
| Chronic kidney disease | 1.43 | (0.91-2.25) | 0.119 |  |  |  |
| Use of ACEi/ARB | 1.05 | (0.69-1.60) | 0.825 |  |  |  |
| Use of β-blockers | 0.85 | (0.39-1.87) | 0.692 | | | |
| Use of MRA | 1.42 | (0.91-2.22) | 0.118 |  |  |  |
| Use of ARNI | 0.58 | (0.35-0.95) | 0.032 | 0.42 | (0.25-0.71) | 0.002 |
| Use of SGLT2i | 0.53 | (0.27-1.03) | 0.062 |  |  |  |
| Use of loop diuretics | 2.83 | (1.45-5.52) | 0.003 |  |  |  |
| Pre-implantation QRSf | 2.98 | (1.67-5.32) | <0.001 | 2.79 | (1.54-5.05) | 0.001 |
| Paced QRS at baseline | 1.03 | (0.62-1.73) | 0.903 |  |  |  |
| Paced QRS width | 1.01 | (1.01-1.02) | 0.002 |  |  |  |
| LVEF at baseline (%) | 0.96 | (0.94-0.99) | 0.006 | 0.97 | (0.94-0.99) | 0.016 |
| LVEF relative increase >10% | 0.53 | (0.34-0.84) | 0.007 |  |  |  |
| LVEF absolute increase >10% | 0.55 | (0.35-0.87) | 0.011 |  |  |  |
| LVEDD at follow-up (mm) | 1.03 | (1.01-1.05) | 0.008 |  |  |  |
| LVEDD reduction | 0.76 | (0.48-1.19) | 0.223 |  |  |  |
| LAD diameter at follow-up (mm) | 1.03 | (1.00-1.05) | 0.022 |  |  |  |
| NYHA I-II at follow-up | 0.29 | (0.18-0.47) | <0.001 | 0.31 | (0.18-0.52) | <0.001 |
| Sinus rhythm at follow-up | 0.43 | (0.27-0.69) | 0.001 | 0.38 | (0.23-0.62) | <0.001 |

ACEi: angiotensin converting enzyme inhibitors; ARB: angiotensin receptor blockers; ARNI: angiotensin receptor/neprilysin inhibitors; CI: confidence interval; CRT: cardiac resynchronization therapy; eGFR: estimated glomerular filtration rate; HR: hazard ratio; LAD: left atrial diameter in 4-chamber view; LVEDD: left ventricular end-diastolic diameter; LVEF: left ventricular ejection fraction; MRA: mineralocorticoid receptor antagonist; NS: non-significant; NYHA: New York Heart Association; QRSf: QRS fragmentation; SGLT2i: sodium-glucose co-transporter 2 inhibitors.

**Figure 2.**
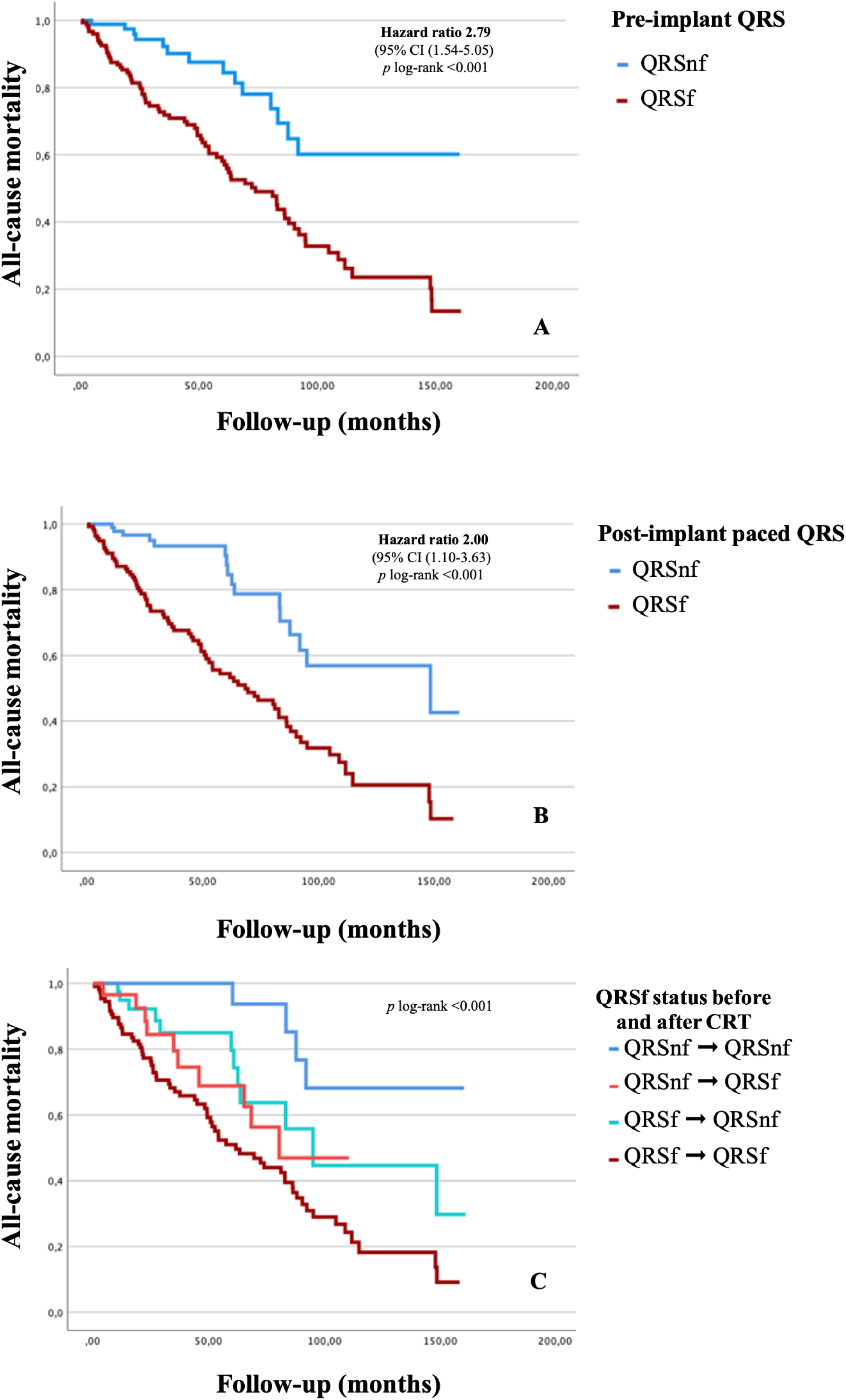
Kaplan-Meier curves for all-cause mortality according to QRSf status before and after CRT. Kaplan-Meier curves for all-cause mortality according to QRSf at baseline (A), after CRT implantation (B), and status before and after CRT (C).

### Hospitalization for HF

A total of 92 patients (37.7%) had at least one hospitalization for HF during the follow-up period. Of these participants, 79 had QRSf before CRT implantation and 78 developed QRSf after implantation. When analyzing the response to CRT in terms of QRSf, 2 of 64 patients with no QRSf before or after CRT were hospitalized for HF, whereas 67 of 110 patients with QRSf before and after CRT were hospitalized for HF. Eleven of 29 patients who developed paced QRSf after CRT were hospitalized for HF, whereas 12 of 28 in whom QRSf resolved after pacing with CRT were hospitalized for HF (Table 2).

Univariate Cox regression analysis revealed that hospitalization for HF events was associated with age, female sex, chronic kidney disease, use of loop diuretics, QRSf before CRT implantation, paced QRS width after CRT, and LVEDD and LAD at follow-up. Only age and pre-implantation QRSf were independently associated with HF hospitalization events on multivariate analysis using Cox regression. Baseline QRSf appeared to be the strongest predictor of HF hospitalization (HR, 3.57; 95% CI [1.94-6.58]) (Table 5). When the influence of post-CRT QRSf instead of pre-implantation QRSf was evaluated on multivariate analysis, it was also a strong predictor of HF hospitalization (Table S4). When analyzing the influence of QRSf status before and after implantation, the multivariate Cox regression survival analysis showed that development of QRSf after CRT implantation (HR, 8.87; 95% CI [1.85-42.50]) and the presence of QRSf before implantation with or without paced QRSf after CRT (HR, 8.21; 95% CI [1.75-38.50]) and HR, 16.70; 95% CI [3.88-71.80], respectively) were independently associated with HF hospitalizations (Table S5). Kaplan-Meier curves comparing native QRSf, paced QRSf, and QRSf status before and after CRT with regard to HF hospitalizations are presented in Figures 3A, 3B, and 3C, respectively.

**Table 5.** Univariate and multivariate analysis using Cox regression for HF hospitalization according to pre-implantation QRSf.

| Hospitalization for HF | Unadjusted analysis |  |  | Adjusted analysis |  |  |
| --- | --- | --- | --- | --- | --- | --- |
|  | HR | 95% CI | P-value | HR | 95% CI | P-value |
| Age (years) | 1.04 | (1.02-1.07) | 0.002 | 1.03 | (1.01-1.06) | 0.020 |
| Sex | 1.73 | (0.98-3.04) | 0.058 |  |  |  |
| Diabetes | 0.95 | (0.62-1.46) | 0.814 |  |  |  |
| Hypertension | 1.37 | (0.86-2.20) | 0.187 |  |  |  |
| Non-ischemic etiology | 1.46 | (0.96-2.21) | 0.078 |  |  |  |
| eGFR (ml) | 0.98 | (0.97-0.99) | <0.001 | 0.98 | (0.97-0.99) | 0.001 |
| Chronic kidney disease | 2.43 | (1.51-3.93) | <0.001 |  |  |  |
| Use of ACEi/ARB | 0.59 | (0.38-0.91) | 0.018 |  |  |  |
| Use of BB | 0.71 | (0.34-1.48) | 0.358 |  |  |  |
| Use of MRA | 1.40 | (0.90-2.16) | 0.133 |  |  |  |
| Use of ARNI | 1.01 | (0.65-1.55) | 0.972 |  |  |  |
| Use of SGLT2i | 1.12 | (0.68-1.83) | 0.657 |  |  |  |
| Use of loop diuretics | 3.62 | (1.80-7.29) | <0.001 |  |  |  |
| Pre-implantation QRSf | 4.33 | (2.39-7.85) | <0.001 | 3.57 | (1.94-6.58) | <0.001 |
| Paced QRS at baseline | 1.33 | (0.82-2.15) | 0.243 |  |  |  |
| Paced QRS width | 1.01 | (1.00-1.02) | 0.008 |  |  |  |
| LVEF at baseline (%) | 0.98 | (0.95-1.00) | 0.083 |  |  |  |
| LVEF relative increase >10% | 0.50 | (0.31-0.80) | 0.005 |  |  |  |
| LVEF absolute increase >10% | 0.50 | (0.31-0.80) | 0.005 |  |  |  |
| LVEDD at follow-up (mm) | 1.04 | (1.02-1.07) | <0.001 |  |  |  |
| LVEDD reduction at follow-up | 0.54 | (0.35-0.86) | 0.009 | 0.54 | (0.33-0.89) | 0.016 |
| LAD at follow-up (mm) | 1.05 | (1.02-1.07) | <0.001 |  |  |  |
| NYHA I-II at follow-up | 0.36 | (0.22-0.60) | <0.001 | 0.39 | (0.23-0.66) | 0.001 |
| Sinus rhythm at follow-up | 0.49 | (0.31-0.76) | 0.002 | 0.57 | (0.36-0.90) | 0.017 |
| ACEi: angiotensin converting enzyme inhibitors; ARB: angiotensin receptor blockers; ARNI: angiotensin receptor/neprilysin inhibitors; CI: confidence interval; CRT: cardiac resynchronization therapy; eGFR: estimated glomerular filtration rate; HR: hazard ratio; LAD: left atrial diameter in 4-chamber view; LVEDD: left ventricular end-diastolic diameter; LVEF: left ventricular ejection fraction; MRA: mineralocorticoid receptor antagonist; NS: non-significant; NYHA: New York Heart Association; QRSf: QRS fragmentation; SGLT2i: sodium-glucose co-transporter 2 inhibitors. |  |  |  |  |  |  |

**Figure 3.**
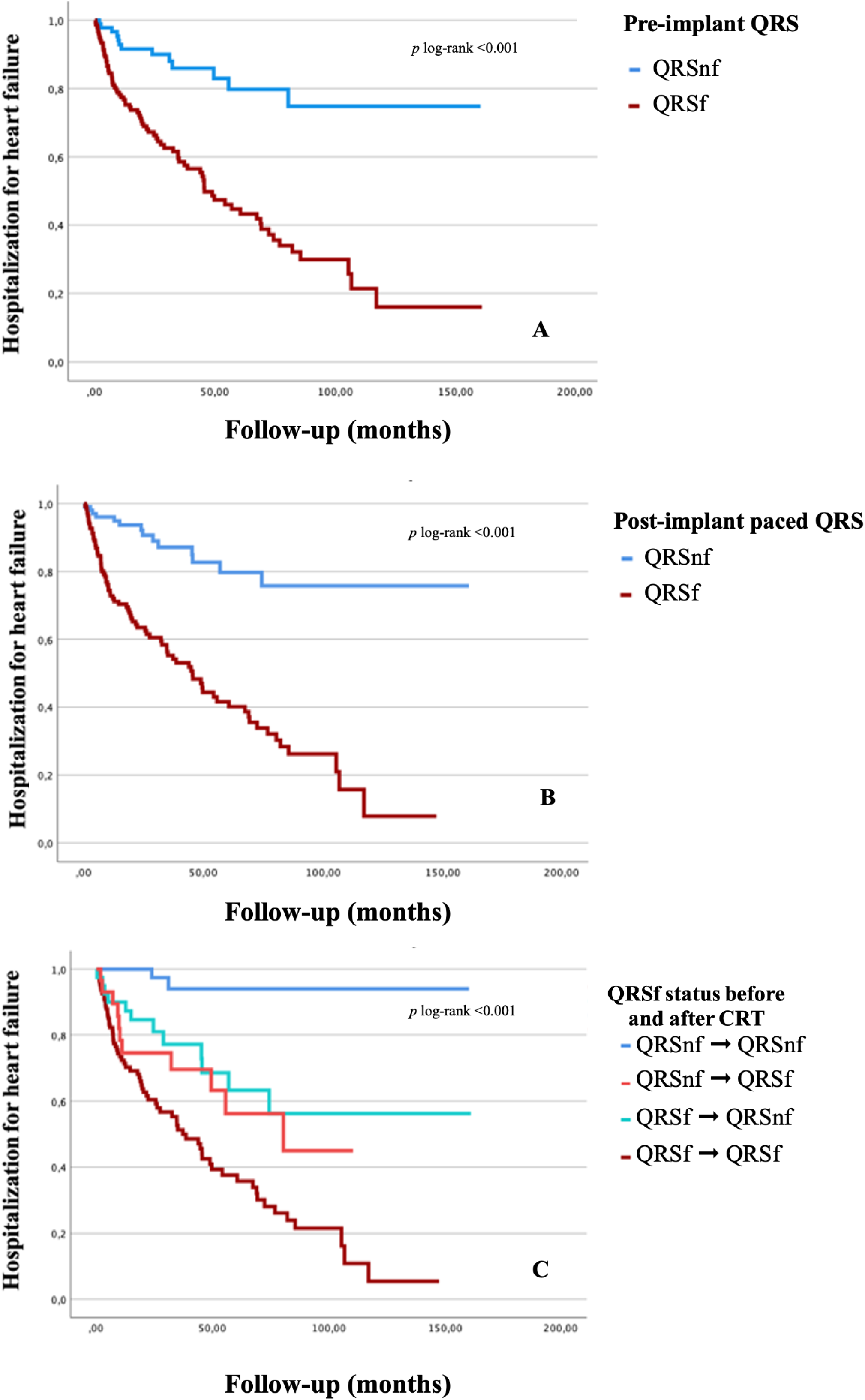
Kaplan-Meier curves for hospitalization for heart failure according to QRSf status before and after CRT. Kaplan-Meier curves for hospitalization for HF according to QRSf at baseline (A), after CRT implantation (B), and status before and after CRT (C).

### Arrhythmic events

A total of 29 patients with CRT-D (16.5%) met the composite endpoint of sustained ventricular tachycardia or appropriate ICD therapy. Twenty-seven participants with pre-implantation QRSf achieved the secondary composite endpoint, while only 3 patients without pre-implantation QRSf experienced AEs. A total of 23 patients with paced QRSf met the secondary composite endpoint. An analysis of the occurrence of AEs in relation to the development, resolution, or persistence of QRSf showed that 23 participants with QRSf before and after CRT implantation experienced this secondary endpoint (Table 3).

AEs were only significantly associated with the presence of pre-implantation QRSf and the use of iSGLT2 as shown on univariate analysis. As shown by multivariate analysis using Cox regression, the presence of pre-implantation QRSf (HR, 6.99; 95% CI [1.54 -31.68]) and use of iSGLT2 remained significantly associated with ventricular arrhythmias or appropriate ICD therapies at long-term follow-up (Table 6). The Cox regression model showed a non-significant trend toward paced QRSf for the prediction of AEs when paced QRSf was included instead of pre-implantation QRSf (Table S6). When analyzing the predictive value of QRSf status before and after CRT, a multivariate survival analysis using Cox regression showed that presence of QRSf before and after CRT-D was an independent predictor of AEs (HR 3.56; 95% CI [1.00-12.70]) (Table S7). Kaplan-Meier analysis revealed significant between-group differences in survival without ventricular arrhythmia events or appropriate ICD therapies based on QRSf status before and after CRT implantation (Figure 4A and 4B) and with respect to resolution, development, or persistence of QRSf after CRT (Figure 4C).

**Table 6.** Univariate and multivariate analysis using Cox regression for arrhythmic events regarding pre-implantation QRSf.

**Title:** Table 6. Univariate and multivariate analysis using Cox regression for arrhythmic
| Arrhythmic event | Unadjusted analysis |  |  | Adjusted analysis |  |  |
| --- | --- | --- | --- | --- | --- | --- |
|  | HR | 95% CI | P-value | HR | 95% CI | P-value |
| Age (years) | 0.98 | (0.95-1.02) | 0.336 |  |  |  |
| Female sex | 0.89 | (0.37-2.19) | 0.800 |  |  |  |
| Diabetes | 0.85 | (0.37-1.93) | 0.680 |  |  |  |
| Hypertension | 1.09 | (0.49-2.42) | 0.836 |  |  |  |
| Non-ischemic etiology | 1.06 | (0.49-2.29) | 0.870 |  |  |  |
| eGFR (ml) | 1.00 | (0.98-1.01) | 0.836 |  |  |  |
| Chronic kidney disease | 1.09 | (0.50-2.40) | 0.823 |  |  |  |
| Use of ACEi/ARB | 0.77 | (0.35-1.68) | 0.498 |  |  |  |
| Use of BB | 0.61 | (0.14-2.78) | 0.512 |  |  |  |
| Use of MRA | 2.09 | (0.88-4.93) | 0.090 |  |  |  |
| Use of ARNI | 1.51 | (0.70-3.27) | 0.280 |  |  |  |
| Use of iSGLT2 | 2.80 | (1.25-6.31) | 0.015 | 2.69 | (1.21-5.99) | 0.017 |
| Use of loop diuretics | 1.09 | (0.44-2.65) | 0.851 |  |  |  |
| Pre-implantation QRSf | 7.10 | (1.57-32.0) | 0.013 | 6.99 | (1.54-31.68) | 0.014 |
| Paced QRS at baseline | 0.64 | (0.18-2.26) | 0.477 |  |  |  |
| Paced QRS width | 1.01 | (0.99-1.03) | 0.205 |  |  |  |
| LVEF at baseline (%) | 1.00 | (0.96-1.05) | 0.824 |  |  |  |
| LVEF relative increase >10% | 0.68 | (0.29-1.60) | 0.357 |  |  |  |
| LVEF absolute increase >10% | 0.66 | (0.28-1.57) | 0.332 |  |  |  |
| LVEDD at follow-up (mm) | 1.04 | (1.00-1.08) | 0.081 |  |  |  |
| LVEDD reduction at follow-up | 0.41 | (0.18-0.94) | 0.036 |  |  |  |
| LAAPD at follow-up (mm) | 1.01 | (0.96-1.05) | 0.803 |  |  |  |
| NYHA I-II at follow-up | 0.91 | (0.28-2.98) | 0.873 |  |  |  |
| Sinus rhythm at follow-up | 1.03 | (0.47-2.25) | 0.937 |  |  |  |
| ACEi: angiotensin converting enzyme inhibitors; ARB: angiotensin receptor blockers; ARNI: angiotensin receptor/neprilysin inhibitors; CI: confidence interval; CRT: cardiac resynchronization therapy; eGFR: estimated glomerular filtration rate; HR: hazard ratio; LAD: left atrial diameter in 4-chamber view; LVEDD: left ventricular end-diastolic diameter; LVEF: left ventricular ejection fraction; MRA: mineralocorticoid receptor antagonist; NS: non-significant; NYHA: New York Heart Association; QRSf: QRS fragmentation; SGLT2i: sodium-glucose co-transporter 2 inhibitors. |  |  |  |  |  |  |

**Figure 4.**
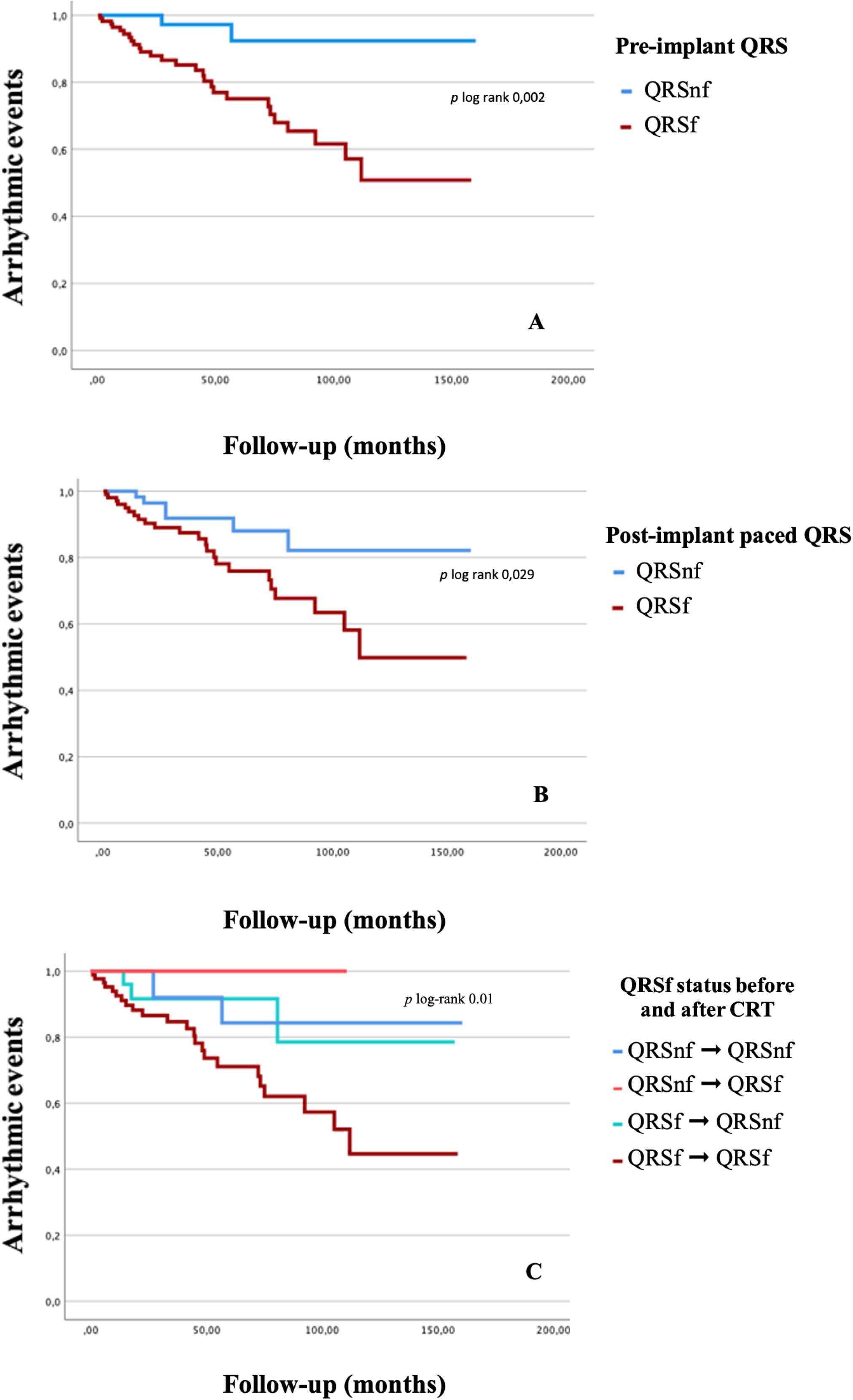
Kaplan-Meier curves for arrhythmic events according to QRSf status before and after CRT. Kaplan-Meier curves for arrhythmic events for patients with QRSf at baseline (A) or after CRT implantation (B), and according to status before and after CRT (C).

## DISCUSSION

In our study, we demonstrated that the presence of QRSf before or after CRT implantation was independently associated with all-cause mortality and HF hospitalization and with a higher incidence of AEs at follow-up in patients undergoing CRT-D. In addition, we observed significant differences in LVEF and LVEDD at follow-up based on the presence of QRSf either before or after implantation. Our results also suggest that QRSf is a dynamic feature, as a small proportion of patients experienced newly developed QRSf or resolution of fragmentation with CRT. Participants in whom QRSf resolved after CRT had a better prognosis than those with persistent QRSf, and patients who developed QRSf after resynchronization had a worse prognosis than those without this finding.

In recent years, QRSf has been studied in several cardiac diseases, particularly ischemic heart disease (IHD) (17,18) and non-ischemic dilated cardiomyopathy (DCM) (7). Das et al (6) suggested that QRSf is a more sensitive indicator of myocardial scar and has a higher negative predictive value than the presence of Q waves alone, and Yooprasert et al (19) showed that fragmentation was associated with myocardial scar as assessed by cardiac magnetic resonance (CMR) imaging. However, this finding remains controversial, as inconsistent results have been published in other studies (20,21). Das et al also reported an increased risk of cardiac events except all-cause mortality in a cohort of 998 patients with IHD (22) and found an association with appropriate ICD shocks in patients with DCM (23). Sha et al (24) concluded that QRSf (QRS <120 ms) was independently associated with all-cause mortality and AEs, and more recently, Marume et al (25) showed an association of QRSf (QRS <120 ms) with all-cause death and MACE but not with the composite arrhythmic events, both in non-ischemic DCM cohorts. A meta-analysis (26) that included 5009 patients with CAD or non-ischemic cardiomyopathy from studies considering QRSf of any width showed that QRSf was associated with an increased risk of sudden cardiac death and all-cause mortality, especially in those patients with LVEF <35% and a width <120 ms.

Previous research suggests that QRSf is of limited significance in patients undergoing CRT. A study involving 233 patients with CRT found no association between adverse left ventricular remodeling, HF events and mortality and the presence QRSf (27). The conclusions of this study may be limited by the high proportion of female participants in the sample, a short follow-up time, and low prevalence of QRSf. However, a small-scale study suggested that the presence of QRSf is a potentially useful marker to identify CRT non-responders (28), and another small study in patients with non-ischemic DCM undergoing CRT showed an increased risk of AEs in patients with post-implantation QRSf (11). Interestingly, the study suggests a possible relationship between lead positioning and the presence of paced QRSf, although the authors performed no imaging studies of scars. Similarly, other small studies (9,10,29) support the hypothesis that QRSf is a predictor of non-response to CRT and further suggest that the resolution of QRSf could be a marker of reverse remodeling.

Since evidence of the prognostic value of QRSf is based on small and heterogeneous studies, we performed an observational analysis to assess the prognostic significance of pre- and post-implantation QRSf in patients with HF with reduced ejection fraction undergoing CRT in terms of all-cause mortality, HF hospitalization, and AEs. Contrary to some of the existing research mentioned above, which excluded wide QRS complexes from the definition of QRSf, we selected this subpopulation with mostly wide QRS complexes in order to investigate the prognostic value of fragmentation independently of QRS width.

Our study includes one of the largest cohorts of patients undergoing CRT to assess the role of QRSf, and the results are consistent with previous evidence regarding hard endpoints such as all-cause mortality and HF hospitalization. QRSf was also a strong predictor of AEs in the subpopulation of patients undergoing CRT-D. Additionally, even though resolution of QRSf has been suggested as a surrogate for responsiveness to CRT, no studies have assessed the prognostic significance of changes in QRS complex fragmentation in terms of adverse outcomes. Despite the scant evidence on QRSf, it could be used as a prognostic marker throughout the follow-up of patients undergoing CRT, and possibly other cardiac conditions.

Although no differences between groups for the prespecified secondary endpoint of a ≥10% increase in LVEF or any improvement of LVEDD (only patients without paced QRSf met the LVEF improvement endpoint), patients without QRSf either before or after CRT implantation seemed to have significantly greater LVEF, smaller LVEDD and higher proportion of NYHA functional class I-II at follow-up compared with those with QRSf. Regarding persistent, newly developed, or resolved QRSf after CRT, patients with QRSf before and after CRT seem to have the worst LVEF and LVEDD values at follow-up.

QRSf is a simple, non-invasive electrocardiographic feature that is widely available and provides useful prognostic information during follow-up. Use of QRSf may be highly relevant for stratifying risk, identifying non-responders, and tailoring the management and follow-up of these patients. Further studies are required to assess the correlation between fragmentation of the QRS complex and cardiac remodeling parameters assessed by CMR and the relevance of changes in serial surface ECG findings over time in various subsets of patients.

### Limitations

Our study has several limitations. First, as QRSf appears to be a dynamic characteristic of the surface ECG that may depend on pacing programming or rhythm disturbances, evaluation of 2 single ECGs before and after CRT implantation may be insufficient to give a precise assessment of how relevant changes are in terms of prognosis. **Furthermore, our study does not assess the significance of the reduction in the number of derivations with QRSf or the change in location of QRSf with CRT-pacing**. Second, this is a single-center retrospective observational study, and some patients were lost to follow-up. Third, we evaluated response to CRT using TTE methods (LVEDD instead of left ventricle end diastolic volume and LVEF) and clinical parameters (NYHA). In addition, prospective studies should analyze response to CRT with CMR index parameters (including LV end-diastolic volume index), which may more accurately evaluate CRT response. Finally, a small proportion of our cohort underwent CRT with left bundle branch pacing (LBBP). Although no statistically significant differences were found between classical CRT and LBBP, likely due to the limited sample size of the latter, this may have an impact on prognosis and response. Therefore, further studies are needed to analyze the effect of LBBP on QRSf and its possible prognostic implications.

## CONCLUSIONS

The presence of QRSf before or after CRT implantation and changes in fragmentation status after implantation are strongly associated with all-cause mortality, hospitalization for HF, and AEs on long-term follow-up. This is the first study to evaluate the significance of pre- and post-implantation QRSf and the prognostic implications of the resolution or development of paced QRSf.

## Data Availability

All data supporting the findings of this study are available within the article and its supplementary materials. Additional data and details can be provided by the corresponding author upon reasonable request.

## Abbreviations

AEs: Arrhythmic events
AV: Atrioventricular
CI: Confidence interval
CMR: Cardiac magnetic resonance
CRT: Cardiac resynchronization therapy
CRT-D: Cardiac resynchronization with a defibrillator
DCM: Dilated cardiomyopathy
HF: Heart failure
HR: Hazard ratio
ICD: Implantable cardioverter defibrillator
IHD: Ischemic heart disease
IQR: Interquartile range
LAAPD: Left atrial anteroposterior diameter
LBBB: Left bundle branch block
LBBP: Left bundle branch pacing
LVEDD: Left ventricle end-diastolic diameter
LVEF: Left ventricle ejection fraction
MPP: Multipoint pacing
NYHA: New York Heart Association
QRSf: QRS fragmentation
SD: Standard deviation
TTE: Transthoracic echocardiography
VV: Interventricular

**Central illustration**

**Title**: QRS fragmentation as a predictor of clinical events in patients undergoing cardiac resynchronization therapy

**Caption:** Two hundred forty-four patients with HF and reduced ejection fraction undergoing CRT were included and presence of QRSf before and after CRT implantation, as well as clinical events at follow up were analysed. In multivariate Cox regression models and Kaplan-Meier curves, QRSf before or after CRT has significant prognostic value for all-cause mortality, HF hospitalization, and AEs, and presence of QRSf after implantation also has prognostic significance.

